# A genome-wide association study of polycystic ovary syndrome identified from electronic health records

**DOI:** 10.1101/2019.12.12.19014761

**Authors:** Yanfei Zhang, Kevin Ho, Jacob M. Keaton, Dustin N. Hartzel, Felix Day, Anne E. Justice, Navya S. Josyula, Sarah A. Pendergrass, Ky’Era Actkins, Lea K. Davis, Digna R. Velez Edwards, Brody Holohan, Andrea Ramirez, Ian B. Stanaway, David R. Crosslin, Gail P. Jarvik, Patrick Sleiman, Hakon Hakonarson, Marc S. Williams, Ming Ta Michael Lee

## Abstract

**Background:** Polycystic ovary syndrome (PCOS) is the most common endocrine disorder affecting women of reproductive age. Previous studies have identified genetic variants associated with PCOS identified by different diagnostic criteria. The Rotterdam Criteria is the broadest and able to identify the most PCOS cases.

**Objectives:** To identify novel associated genetic variants, we extracted PCOS cases and controls from the electronic health records (EHR) based on the Rotterdam Criteria and performed a genome-wide association study (GWAS).

**Study Design:** We developed a PCOS phenotyping algorithm based on the Rotterdam criteria and applied it to three EHR-linked biobanks to identify cases and controls for genetic study. In discovery phase, we performed individual GWAS using the Geisinger’s MyCode and the eMERGE cohorts, which were then meta-analyzed. We attempted validation of the significantly association loci (P<1×10^−6^) in the BioVU cohort. All association analyses used logistic regression, assuming an additive genetic model, and adjusted for principal components to control for population stratification. An inverse-variance fixed effect model was adopted for meta-analyses. Additionally, we examined the top variants to evaluate their associations with each criterion in the phenotyping algorithm. We used STRING to identify protein-protein interaction network.

**Results:** We identified 2,995 PCOS cases and 53,599 controls in total (2,742cases and 51,438 controls from the discovery phase; 253 cases and 2,161 controls in the validation phase). GWAS identified one novel genome-wide significant variant rs17186366 (OR=1.37 [1.23,1.54], P=2.8×10^−8^) located near *SOD2*. Additionally, two loci with suggestive association were also identified: rs113168128 (OR=1.72 [1.42,2.10], P=5.2 x10^−8^), an intronic variant of *ERBB4* that is independent from the previously published variants, and rs144248326 (OR=2.13 [1.52,2.86], P=8.45×10^−7^), a novel intronic variant in *WWTR1*. In the further association tests of the top 3 SNPs with each criterion in the PCOS algorithm, we found that rs17186366 was associated with polycystic and hyperandrogenism, while rs11316812 and rs144248326 were mainly associated with oligomenorrhea or infertility. Besides ERBB4, we also validated the association with *DENND1A1*.

**Conclusion:** Through a discovery-validation GWAS on PCOS cases and controls identified from EHR using an algorithm based on Rotterdam criteria, we identified and validated a novel association with variants within *ERBB4*. We also identified novel associations nearby *SOD2* and *WWTR1*. These results suggest the eGFR and Hippo pathways in the disease etiology. With previously identified PCOS-associated loci *YAP1*, the *ERBB4-YAP1-WWTR1* network implicates the epidermal growth factor receptor and the Hippo pathway in the multifactorial etiology of PCOS.

## Introduction

Polycystic ovary syndrome (PCOS) is the most common endocrine disorder that affects women of reproductive age ^1^. PCOS is characterized by the three main features: dysregulation of the menstrual cycle; elevated levels of androgenic hormones; and multiple cysts of the ovaries from which the name of the condition is derived. Other features include hirsutism in a “male” pattern, acne, increased skin pigment sometimes associated with skin tags, and weight gain. Three criteria to identify women with PCOS have been proposed: the National Institutes of Health (NIH) Criteria, the Rotterdam Criteria, and the Androgen Excess and PCOS Society (AE-PCOS) criteria. The NIH criteria requires both hyperandrogenism and oligomenorrhea ^2^; the Rotterdam criteria requires at least two of the three phenotypes: hyperandrogenism, oligo-ovulation, and polycystic ovaries ^3^; and the AE-PCOS requires both hyperandrogenism and ovarian disfunction ^4^. The Rotterdam criteria is more inclusive than the other two which increases its sensitivity, thus, the prevalence of PCOS estimated by the Rotterdam criteria is 15-20% compared to the 7-12% generated by the other two criteria ^5, 6^.

Heritability estimated for PCOS ranges 38-71% by twin studies ^7^, with a polygenic genetic architecture and complex inheritance pattern ^8, 9^. Recent large-scale genome-wide association studies (GWAS) have identified 19 loci associated with PCOS in women with European or East Asian ancestries, including *ERBB4, YAP1* and *DENND1A* that were replicated in European and Asian ancestral groups, providing additional evidence for the polygenic architecture of PCOS ^10-16^. These studies adopted different diagnosis criteria, including PCOS cases diagnosed based on NIH or Rotterdam criteria, or self-reported information. Shared genetic architecture for PCOS using the different diagnosis criteria or self-reported were also identified by genetic correlation analyses ^13, 16^.

The healthcare system-based biobanks with genetic data linked to the electronic health record (EHR) data enable new opportunities for genomic discovery research ^17^. Examples include Geisinger’s MyCode^®^ Community Health Initiative (MyCode) ^18^, BioVU at Vanderbilt University ^19, 20^, and the <u>e</u>lectronic <u>ME</u>dical <u>R</u>ecords and <u>GE</u>nomics (eMERGE) Network, a nationwide consortium of multiple medical institutions that link DNA biobanks to EHRs ^21^. These multidimensional data are important resources for development of phenotype algorithms, genetic discoveries, and clinical implementation ^22-24^. Phenotyping algorithms to identify cases and controls for various diseases from EHR have been developed ^25^. Such approaches are critical for genetic studies as they integrate data from different EHR systems derived using the same phenotype definition that has been rigorously evaluated to define the performance characteristics in order to reduce case selection bias and heterogeneity among different studies.

In this study, we aim to develop an EHR algorithm for PCOS based on the Rotterdam criteria to identified PCOS cases and controls in multiple cohort and perform a GWAS to identify genetic variants associated with PCOS.

## Cohort and Methods

### Cohorts

The discovery cohorts were identified from the Geisinger MyCode Community Health Initiative (MyCode) Phase I ∼ II and eMERGE Phase III. All MyCode participants provide written consent allowing their clinical and genomic data to be used for health-related research ^18, 26^. The eMERGE Phase III includes 83,717 individuals recruited from 12 study sites with demographics, diagnosis information based on ICD codes, and genotyping data ^24^. The replication cohort was selected from BioVU, Vanderbilt University’s EHR-linked biorepository ^19, 20^. This study was waived for a standard institutional review board (IRB) review based on the use of deidentified EHR and genetic data from all sites. We received approval from the Geisinger MyCode Governing Board, the eMERGE coordinating center and the BioVU Review Committee and IRB to conduct this genetic study. Since both Geisinger and Vanderbilt are eMERGE sites, participants in MyCode and BioVU who were included in the eMERGE data were excluded from the site-specific analysis to avoid double-counting.

### PCOS EHR algorithm based on the Rotterdam Criteria

**Figure 1** illustrates the sample selection and analytic strategy of this study. The Geisinger PCOS EHR algorithm based on the Rotterdam diagnosis criteria was first developed to identify PCOS cases and controls from the EHR data. The three criteria that were used in the algorithm to represent different aspects of PCOS are: 1) Polycystic (C1): having diagnosis codes of polycystic ovarian syndrome and/or polycystic ovaries; 2) Hyperandrogenic (C2): having diagnosis codes for hyperandrogenism or hyperandrogenism-related clinical signs or hyperandrogenemia determined by testosterone measurements; 3) Reproductive (C3): having diagnosis codes for oligomenorrhea, amenorrhea, infertility and oligo-or anovulation. **Supplementary Tabel 1** provides details and the inclusion and exclusion ICD codes and laboratory tests for each criterion. PCOS cases were patients that met at least 2 of the 3 criteria with an index age between 18 to 45. Controls were those who did not have any components of the three criteria, and whose current age was older than the median age of the cases (38 years in this study) to increase the specificity for the controls. This algorithm was then applied to the Geisinger and eMERGE cohorts for the discovery GWAS.

### Discovery GWAS and meta-analyses

MyCode Phase I and Phase II samples were genotyped and imputed to HRC.r1-1 EUR reference genome (GRCh37 build) separately using the Michigan Imputation Server as previously described ^27^. Variants with imputation info score > 0.7 were included for analyses. eMERGE samples were genotyped at each study site and imputed to HRC.r1-1 EUR reference genome in multiple batches using the Michigan Imputation Server. Data were processed centrally and harmonized as previously described ^24^. Variants with average info score >0.3 were included. Samples with a genotyping rate below 95% were excluded. SNPs with a <99% call rate, minor allele frequency (MAF) of <1% and a significant deviation from the Hardy-Weinberg equilibrium (P<1×10^−7^) were removed from analyses. One of the paired individuals with first-or second-degree relatedness were removed. Finally, there were 7,595,111 SNPs, 6,747,339 SNPs, and 5,648,769 SNPs from MyCode Phase I (1,141 cases and 18,788 controls), MyCode Phase II (594 cases and 9,024 controls), and eMERGE III (1,007 cases and 23,626 controls) included for GWAS.

**Figure 1:**
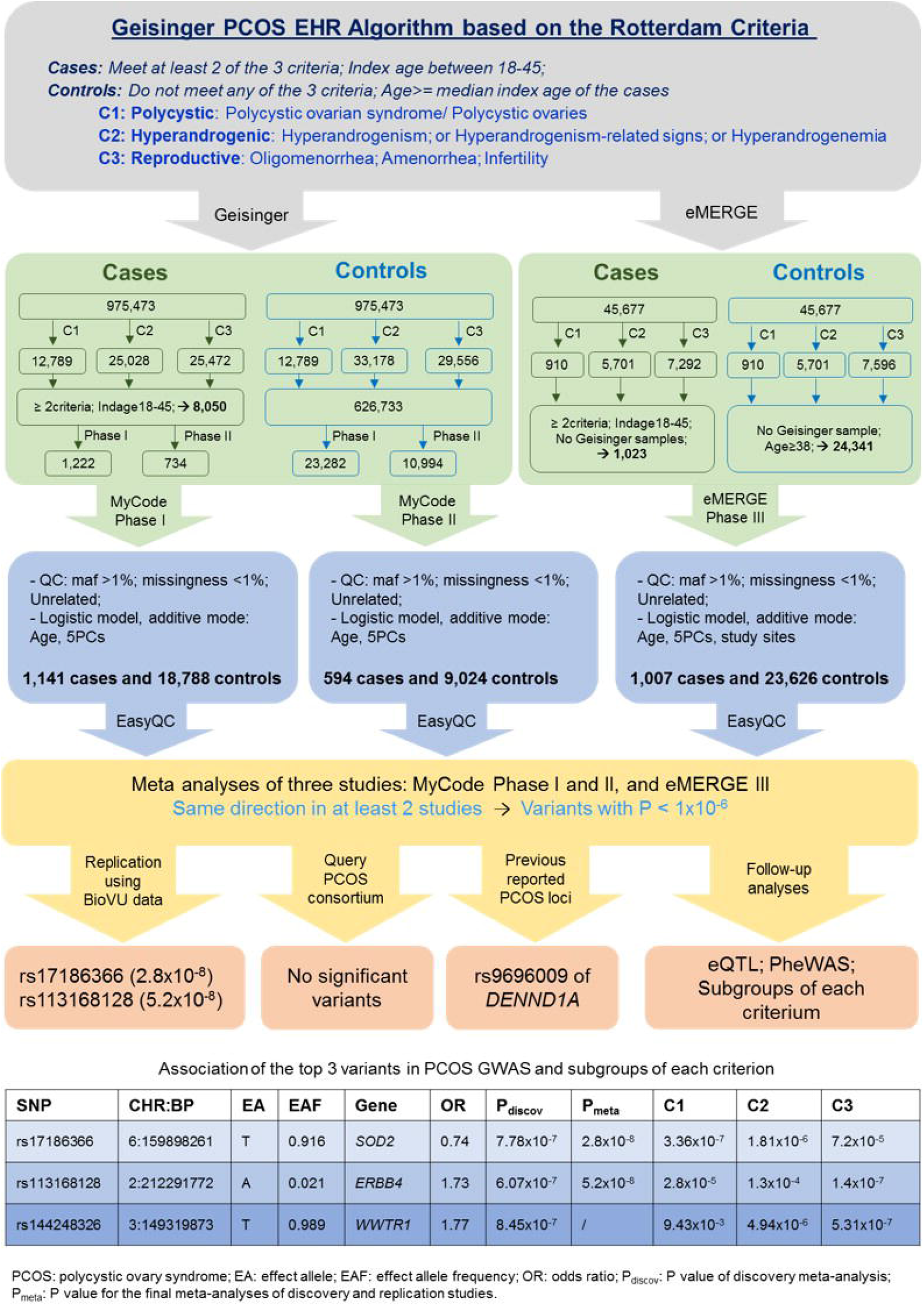
Flowchart for study design. **The** PCOS algorithm was first developed to identify cases and controls from the EHR based on Rotterdam criteria, and then applied to Geisinger patients, and the eMERGE cohort. Case-control GWAS were then conducted for the three cohorts with genetics data followed by fixed effect inverse-variance meta-analyses. Variants with P<1e-6 were then validated in BioVU samples using the same phenotype algorithm and genetics analyses protocol for in meta-analysis and were queried in summary statistics from the PCOS consortium. Association with each criterion in the PCOS algorithm were further tested for these variants.

For study-specific GWAS, we used fixed effects logistic regression, assuming an additive genetic model, adjusted for index age and the first six principal components (PCs) to account for population stratification for the MyCode Phase I and II cohorts; additionally, we adjusted for the eMERGE III study sites. EasyQC ^28^ was employed to harmonize the alleles and data format for GWAS summary statistics from discovery studies before performing a fixed effect inverse variance weighted meta-analysis using METAL ^29^. PLINK 1.9 ^30^ was used to calculate PCs, relatedness and to perform GWAS.

### Replication for the top variants

Top associated variants with P<1×10^−6^ from discovery meta-analysis were further evaluated in an independent PCOS cohort identified based on the same algorithm from BioVU. We identified 253 cases and 2161 controls. Genotypes were generated using the Illumina Infinium Expanded Multi-Ethnic Genotyping Array. The same imputation, quality control measures and association protocols were applied for the replication study. We also queried the summary statistics of the meta-analyses from the PCOS consortium (without the 23andMe data) for the associations of these top variants ^16^. Criteria for replication is P<0.05, directionally consistent in the replication GWAS, or P<5×10^−8^ in the combined meta-analyses.

### Power calculation

We evaluated the power for our study conservatively assuming a significance level of P<5×10^−8^ for GWAS, and a PCOS prevalence of 8%. Given the current PCOS case number of 2995, we have 80% power to identify an associated variant with a MAF of 1% and an OR (odds ratio) > 2.01; or a MAF at 2% and an OR > 1.68; or a MAF > 8% and an OR > 1.34.

### Functional genomics exploration

The Variant Effect Predictor was used for variant annotation ^31^. The Functional Mapping and Annotation was used in the default setting to generate independent loci and associated pathways ^32^. Open Targets Genetics is an online portal used in this study to query the associated genes, phenome-wide association studies (PheWAS), and the expression quantitative trait loci (eQTLs) of the top associated variants ^33^. The PheWAS data in Open Target Genetics includes the results from UK Biobank GWAS and the GWAS catalog. STRING was used to identify the protein-protein interaction network.

## Results

### Identification and characterization of PCOS cases and controls from EHR data

**Figure S1** illustrates the details of sample ascertainment using the Rotterdam-based algorithm in the Geisinger and eMERGE cohorts. Only non-Hispanic whites were included in the MyCode and BioVU samples; All races were included in the eMERGE samples, 75% were of European American, 17% were of African American and 8% were other race/ethnicity (**Supplementary tabel 2**). **Table 1** summarizes the numbers and characteristics of the identified cases and controls. The proportion of patients with polycystic ovaries was around 40% of the PCOS cases identified in the eMERGE and BioVU data; while this number is over 88% in the Geisinger MyCode Phase I and II data. The Geisinger cohorts also have lower hyperandrogenic features than eMERGE and BioVU cohorts. Over 90% of the patients had reproductive issues in all the three cohorts. Cases showed higher BMI than controls in the MyCode and BioVU samples but not in the eMERGE samples.

### Discovery and replication of the genetic variants associated with the risk of PCOS

Twenty independent loci were identified with P<1×10^−5^ in the discovery meta-analysis of the MyCode and eMERGE cohorts (**Supplementary tabel 3**). Manhattan plots for the meta-analysis and the three discovery studies are shown in **Figure 2A** and **Figure S2**. Variants with P<1×10^−6^ were then examined in an independent cohort identified using the same algorithm from BioVU. **Figure 2B** lists the association of the top three independent SNPs in the discovery and replication cohorts. rs17186366, a novel association located in the promotor flanking region near SOD2 and *LOC101929142* reached genome-wide significance (OR=1.37 [1.23,1.54], P=2.8×10^−8^) in the combined meta-analyses of discovery and replication. We also identified an intronic variant of *ERBB4*, rs113168128, with near genome-wide significance (OR=1.72 [1.42,2.10], P=5.2 x10^−8^). This SNP is independent from the previously reported *ERBB4* variant rs2178575 (r^2^ = 0.001) ^16^. A low-frequency intronic variant of *WWTR1* rs144248326 (MAF = 1.01%), was identified and was also close to genome-wide significance level (OR=2.13 [1.52,2.86], P=8.45×10^−7^). The regional association plots for these three *loci* are shown in **Figure S3**. We also examined the top associations for African Americans in the eMERGE datasets. Only rs113168128 in *ERBB4* passed the standard quality check. This variant has higher MAF and a slightly smaller OR with nominal significance (MAF=0.063, OR=1.64, P=0.0106).

**Table 1.**
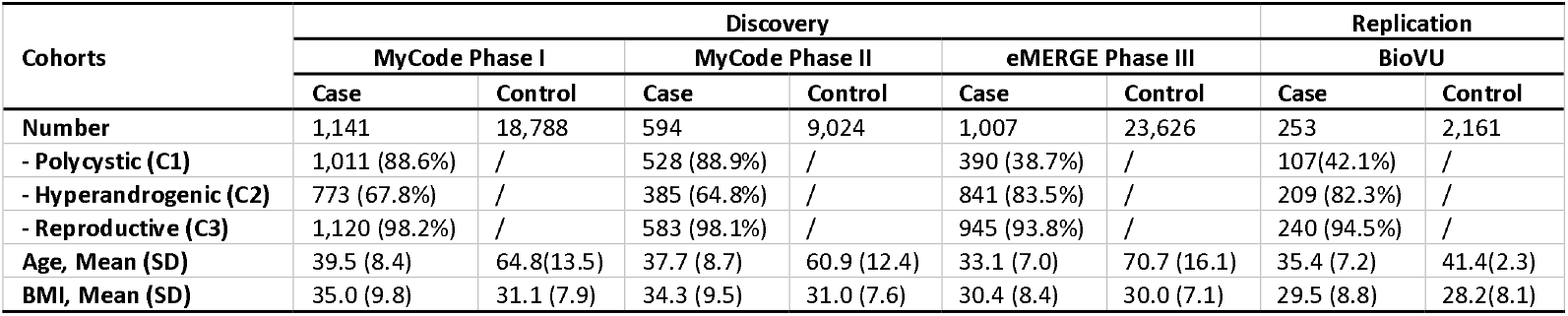
Characteristics of PCOS cases and controls from discovery and replication cohorts.

| Cohorts | Discovery |  |  |  |  |  | Replication |  |
| --- | --- | --- | --- | --- | --- | --- | --- | --- |
|  | MyCode Phase I |  | MyCode Phase II |  | eMERGE Phase III |  | BioVU |  |
|  | Case | Control | Case | Control | Case | Control | Case | Control |
| **Number** | 1,141 | 18,788 | 594 | 9,024 | 1,007 | 23,626 | 253 | 2,161 |
| - Polycystic (C1) | 1,011 (88.6%) | / | 528 (88.9%) | / | 390 (38.7%) | / | 107(42.1%) | / |
| - Hyperandrogenic (C2) | 773 (67.8%) | / | 385 (64.8%) | / | 841 (83.5%) | / | 209 (82.3%) | / |
| - Reproductive (C3) | 1,120 (98.2%) | / | 583 (98.1%) | / | 945 (93.8%) | / | 240 (94.5%) | / |
| **Age, Mean (SD)** | 39.5 (8.4) | 64.8(13.5) | 37.7 (8.7) | 60.9 (12.4) | 33.1 (7.0) | 70.7 (16.1) | 35.4 (7.2) | 41.4(2.3) |
| **BMI, Mean (SD)** | 35.0 (9.8) | 31.1 (7.9) | 34.3 (9.5) | 31.0 (7.6) | 30.4 (8.4) | 30.0 (7.1) | 29.5 (8.8) | 28.2(8.1) |

**Figure 2:**
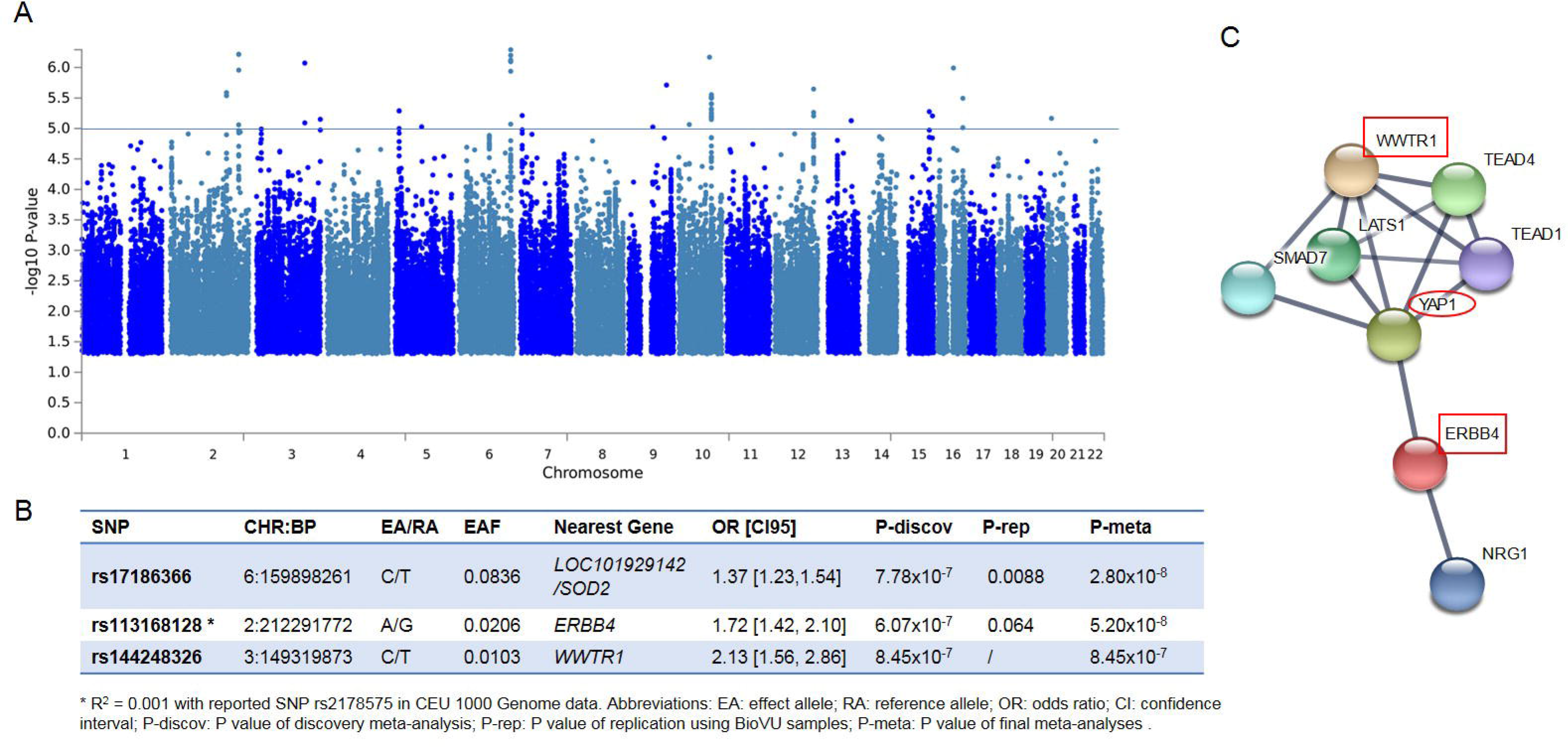
GWAS analyses of PCOS and functional evaluation. (A) Manhattan plot for the meta-analysis of the Geisinger MyCode Phase I, II and the eMERGE Phase III cohorts. (B) The top 3 associated variants with P value < 1e-6 in discovery meta-analysis and their replication and final meta-analysis. (C) The protein-protein interaction (PPI) network for ERBB4, WWTR1 and YAP1 using STRING. Only high confidence interactions were shown (confidence score >=0.7).

We did not observe significant associations of these variants in the PCOS consortium meta-analyses ^16^. We also examined the associations of previously reported PCOS loci in our meta-analyses result. Only variants in *DENND1A1* (rs9696009, rs10818854, rs10986105) were replicated with the same direction and similar effect size (P<0.05; **Supplementary tabel 4**).

The functional genomics exploration found rs17186366 near SOD2 associated with menarche (P=6.6×10^−5^), rs113168128 in ERBB4 associated with depressed affect (P=2.1×10^−5^) ^34^ (**Supplementary tabel 5**). All of the top three SNPs were found to be associated with phenotypes related to the nervous system, or to mental or behavioral disorders (**Supplementary tabel 5**). None of these SNPs were found to be eQTLs in any tissue. The protein-protein interaction network found both ERBB4 and WWTR1 interact with YAP1 (Figure 2C), which also associated with PCOS in both European and Han Chinese ^12, 13, 16^.

### Association of the top three SNPs with each PCOS criterion

**Table 2** summarized the association results for the top three variants with each of the three criteria in the PCOS algorithm that represent the different aspects of PCOS based on the Rotterdam criteria. rs17186366 strongly associated with the polycystic and hyperandrogenic traits, while the other two SNPs in *ERBB4* and *WWTR1* are mainly associated with the reproductive trait as the more significant association P values and larger effect sizes are observed for the variants and the corresponding traits (**Table 2**).

## Discussion

### Principal findings

In this study, we developed an EHR algorithm based on the Rotterdam criteria for PCOS and identified cases and controls from three biobank cohorts. Through a discovery-validation GWAS, we identified rs17186366 near *SOD2*, a novel genome-wide significant signal associated with PCOS. We validated the association of previously reported genes *ERBB4* and *DENND1A1*, with rs113168128 being an independent signal in *ERBB4*. We also identified rs144248326, an intronic variant of *WWTR1*, as a novel signal close to genome-wide significance level. In further association tests of the top 3 SNPs with each criterion in the PCOS algorithm, we found that rs17186366 was associated with polycystic and hyperandrogenism, while rs11316812 and rs144248326 were strongly associated with oligomenorrhea or infertility. The top SNPs are associated with traits related to the nervous system and mental/behavior disorders in the PheWAS queries.

**Table 2:** Association of the top 3 SNPs with each PCOS criterion.

| SNP | Criterion | MyCode Phase I |  | MyCode Phase II |  | eMERGE III |  | Meta |  |
| --- | --- | --- | --- | --- | --- | --- | --- | --- | --- |
|  |  | OR | P | OR | P | OR | P | OR [CI95] | P |
| rs17186366<br>*SOD2* | Polycystic | 1.44 | 0.0004 | 1.54 | 0.0026 | 1.38 | 0.0295 | 1.45[1.25,1.67] | 3.36x10<sup>-7</sup> |
|  | Hyperandrogenic | 1.49 | 0.0003 | 1.48 | 0.0163 | 1.27 | 0.0258 | 1.39[1.21,1.59] | 1.81x10<sup>-6</sup> |
|  | Reproductive | 1.31 | 0.0052 | 1.26 | 0.0992 | 1.27 | 0.0213 | 1.28[1.13,1.45] | 7.23x10<sup>-5</sup> |
| rs113168128<br>*ERBB4* | Polycystic | 2.08 | 0.0004 | 2.19 | 0.0148 | 1.34 | 0.2024 | 1.79[1.36,2.35] | 2.85x10<sup>-5</sup> |
|  | Hyperandrogenic | 1.39 | 0.1818 | 2.27 | 0.0186 | 1.59 | 0.0033 | 1.60[1.26,2.04] | 1.31x10<sup>-4</sup> |
|  | Reproductive | 1.88 | 0.0013 | 2.32 | 0.0040 | 1.62 | 0.0012 | 1.79[1.44,2.22] | 1.40x10<sup>-7</sup> |
| rs144248326<br>*WWTR1* | Polycystic | 1.91 | 0.0276 | 2.71 | 0.0240 | 0.91 | 0.8343 | 1.74[1.15,2.65] | 9.43x10<sup>-3</sup> |
|  | Hyperandrogenic | 1.90 | 0.0406 | 2.50 | 0.0625 | 2.23 | 0.0002 | 2.16[1.55,3.00] | 4.94x10<sup>-6</sup> |
|  | Reproductive | 2.18 | 0.0032 | 3.49 | 0.0011 | 1.87 | 0.0052 | 2.19[1.61,2.97] | 5.31x10<sup>-7</sup> |

## Results and Implications

During case identification, we observed similar proportions in each criterion used in the algorithm for the eMERGE and BioVU data, but different from the MyCode data. Around 40% of the patients had polycystic ovaries in the eMERGE and BioVU cohorts, versus 88% in the Geisinger cohort. This may be due to the integration of information from the patients’ problem list at Geisinger or a difference in clinical practice between the systems.

We identified a novel genome-wide significant variant rs17186366 near SOD2, which was associated with the polycystic ovaries and hyperandrogenism. SOD2 encodes Superoxide Dismutase 2, a mitochondrial protein which converts superoxide byproducts of oxidative phosphorylation to hydrogen peroxide and diatomic oxygen. Recently, A16V (rs4880) in SOD2 was found to be associated with PCOS, serum luteinizing hormone (LH) levels, and the ratio of LH to follicle-stimulating hormone in Han-Chinese women ^35^. rs17186366 was also found to be associated with menarche in the UKB GWAS results. One retrospective study showed early or late menarche were more likely to be seen in women with PCOS ^36^.

The association of *ERBB4* with PCOS has been validated in both European and Han-Chinese ^13, 16, 37^. In this study, we identified an intronic variant rs11316812 of *ERBB4* that is not in LD with the known variants at close to genome-wide significant level (P=5.2×10^−8^). ERBB4, also known as human epidermal growth factor receptor 4 (HER4), belongs to the EGFR family which includes ERBB1, ERBB2/HER2 and ERBB3/HER3. Other than PCOS, variants of *ERBB4* have been associated with ovarian cancer ^38^ and schizophrenia ^39^. ERBB4 can be stimulated by its ligands and activate the EGFR signaling, which is critical for LH-induced steroidogenesis which promotes follicular maturation ^40, 41^. ERBB4 signaling is also involved in luteal growth ^42^. ERBB4 is highly expressed in the nerves system according to GTEx datasets, including in the hypothalamus and pituitary, the two important organs in the hypothalamus-pituitary-ovary-adrenal (HPOA) axis. These findings suggest a disturbance in the control mechanisms of the HPOA axis in PCOS.

*WWTR1*, the third gene with strong association, encodes for TAZ (transcriptional co-activator with PDZ-binding motif). TAZ also contains a WW domain as the Yes-associated protein (YAP1) ^43^ and is another gene that was associated with PCOS ^12, 13, 16^. WWTR1 and YAP1 are two key molecules of the Hippo signaling pathway, and their expression were significantly altered in PCOS tissues ^44^. Insulin resistance affects 50-70% of women with PCOS ^45^. WWTR1 and YAP1 can also regulate insulin resistance. The inhibition of WWTR1/YAP1 in combination with metformin can completely inhibit the effect of insulin ^46^. Our findings provide a possible genetic link between PCOS and the Hippo pathway, suggesting potential pharmaceutical Hippo-targeted interventions for treatment of PCOS with insulin resistance. Interestingly, ERBB4 can also interact with WW domains in YAP1 through the PPxY motif ^47^. The ERBB4-YAP1-WWTR1 interaction network indicates the Hippo and EGFR signaling contribute to the multifactorial etiology of PCOS.

Epidemiologic studies found that women with PCOS have a higher risk for depression, bipolar disorder, anxiety, and eating disorders ^45^. In our study, we found moderate associations for the top three variants with mental/behavioral disorders including depression. When queried, the PheWAS and GWAS catalog through Open Targets Genetics, suggests shared genetic predisposition between PCOS and mental disorders. Also, the differences in association patterns across the PCOS criteria likely reflect the complex biology of PCOS and support the epidemiological observations of heterogeneity in the phenotype.

### Strengths and Limitations

The major strength of this study is the same phenotyping algorithm was applied through different systems to ensure the homogeneity. Our PCOS algorithm was based on the Rotterdam criteria and thus was broader than the NIH or AE-PCOS criteria, enabling us to identify more cases than the ICD code-based method. This would improve the number of cases and avoid the cases that would be identified as controls otherwise, thus increases the specificity of the study. However, the algorithm assumes the same evaluation and coding practices at each healthcare system and its sensitivity and specificity for case identification was not validated by chart review. A potential limitation is case definition of testosterone laboratory measurement (criteria2c) was unavailable in the eMERGE data. However, based on MyCode, all the patients identified by this criterion also met the other two criteria. Thus, absence of this information may not affect the final total case number. Also, the EHR data on luteinizing hormone and follicle-stimulating hormone serum levels were limited thus we were not able to test the associations with the top variants. The sample size is not large enough to identify variants with small effect or low minor allele frequency, especially because PCOS is a heterogenous disorder with complex etiology and combinations of clinical symptoms. For our study, we have about 80% power for the top 3 variants. The study cohorts are mainly European-decent individuals, with a very small proportion of African Americans and other race/ethnic groups. Although the variant in *ERBB4* showed the same direction of effects and nominal significance (P<0.05) in the African American, it has a higher minor allele frequency than in the European population. Future studies focusing on minorities are necessary.

## Conclusions

Through a discovery-validation GWAS on PCOS cases and controls identified from EHR using an algorithm based on Rotterdam criteria, we validated the association with *ERBB4*. We also identified novel association with SOD2 and *WWTR1*. Our findings highlighted the role of EGFR and Hippo signaling in the disturbance of metabolic and HPOA axis in PCOS etiology.

## Data Availability

Summary data is available provided collaboration with Geisinger.

## Acknowledgements

The authors thank Christina M. Yule and Sara J. Kwiecien at Geisinger, and Brittany City at the eMERGE network coordinating center. The authors express their gratitude to Drs. Cecilia Lindgren, John Perry, and Corrine Kolka Welt at the International PCOS Consortium for their support. The authors would like to thank Ilene Ladd for English editing.

## Authors contribution

YZ, KH, AJ, and ML designed the study; KH and DH developed the algorithm and applied to Geisinger EHR; YZ performed the phenotyping for eMERGE data and discovery GWAS; Jacob performed the phenotyping and replication using BioVU data; FD extracted the PCOS consortium data; YZ drafted the manuscript; KH, AJ, ML, SP and MSW did critical review; NJ, LD, AR, GJ, IS, DVE, PS, EA and BH contributed the eMERGE data and provided critical review; All authors approved the final manuscript.

## Funding disclosure

MyCode^®^ was funded by Geisinger and Regeneron Genomics Center; the eMERGE III was funded by NIH U01HG8679 (Geisinger Clinic). The funding sources was not involved in the interpretation of the result or which journal to submit.

References

